# Subclinical left ventricular disfunction in obese children: are we late?

**DOI:** 10.1101/2020.10.10.20195743

**Authors:** B Riaño-Mendez, M Ruiz del Campo, P Garcia-Navas, CM Amiama-Perez de Villarreal, MY Ruiz del Prado

## Abstract

**Aim:** Longitudinal global strain (LGS) is reduced in obese patients with normal ejection fraction of the left ventricle. TAPSE/PAPS ratio, recently described, may be a step forward a more efficient RV function evaluation.

There are still few publications in the application of these methods in pediatric patients.

**Methods:** This case-control study compared 104 children aged 5-18 years between October 2017 and February 2019, 52 obese children with body mass index (BMI) > +2 SD, and 52 matched controls.

They were screened for other cardiovascular risk factors like insulin resistance or hypercholesterolemia. A complete echocardiography including standard and functional parameters was performed.

**Results:** We found that obese children presented poor systolic function (LGS −15,90 ± 3,84 %) in comparison with non-obese children (−19,44 ± 5,75 %, p=0,001). LGS correlated positively with body mass index (BMI).

Standard echocardiography also revealed cardiomegaly and hypertrophy.

TAPSE/PASP ratio correlated negatively with triglycerides levels (β −0,402, p=0,014). Diastolic function was poor in those with HOMA-IR (β −0,375, p=0,016) and hypertriglyceridemia (β −0,375 p=0,024).

**Conclusion:** We think that is necessary to perform a standarised cardiovascular evaluation in obese children for early identification of subclinical dysfunction especially in those with insulin resistance and dyslipidemia

## Introduction

The prevalence of overweight among children in developed and developing countries is continuing to increase^1^. Nearly 40% of paediatric population in our midst are either obese or overweight^2^. One of the most important efforts in public health nowadays in the first world is obesity in children and adolescents. Obese children may be at risk for consequences, both short-term health and long-term tracking of obesity to adulthood^3^.

Intense correspondences between obesity and cardiovascular disease (CVD) have been shown in many studies. In fact, obesity has been considered an independent risk factor for CVD and CVD risks in obese children. Indeed, literature has described the relationship between BMI in adolescence and all-cause mortality^4^.

At present, the secondary and tertiary prevention measures are the most important points to fight against cardiovascular desease^5^. However, efforts to analyse modifiable risk factors present in children and adults over a long time have not been a priority to date^2^.

Obesity in children and adolescents is related with an amount of cardiovascular changes that are interlinked to increased cardiovascular morbidity and mortality in adulthood. Children exhibit early signs of cardiovascular dysfunction as a result of their excess adiposity as recent studies have shown, frequently independent of other obesity-related comorbidities such as insulin resistance, hypertension or dyslipemia. Clinical testimony to date suggests that the cardiovascular injure seen in middle-aged adults is also happening in obese children^6^, particularly in those with a central fat distribution.

The major obesity-induced changes described are dilatation of left ventricular structures, left ventricular (LV) hypertrophy, systolic and diastolic dysfunction, heart failure. Many studies have demonstrated that an appropriate detection and treatment of these cardiovascular damages in early phases is important in order to slow down and reverse the process in high-risk pediatric populations ^7^.

New echocardiographic procedures are developed to accomplish accurate evaluation of myocardial deformation (both segmental and global). The newly released development is the two-dimensional speckle-tracking echocardiography (2D STE), a routine for assessing myocardial strain and measure deformation function^8^.

Two-dimensional speckle tracking imaging is a suitable method for assessing ventricular function and identifying sub-clinical dysfunction. It provides information which is not available with any of the echocardiographic parameters used at the present time. STE provides an impartial quantification of myocardial deformation and left ventricular systolic and diastolic dynamics. One of this value is that this technique is angle-independent. Results regarding the viability, exactitude and clinical application of speckle-tracking echocardiography are quickly accumulating^9^. In literature, there are still few publications in the application of this method in pediatric population.

## Methods

### Study design

The aim of the study was to analyse LV function using 2D speckle tracking in obese pediatric patients referred from the Endocrinology consult with BMI >+2 DS, and to contrast their data with non-obese patients with structurally normal hearts and same age and sex.

We used preceding researches where LV Doppler-derived strain was contrasted between obese and non-obese children for sample size estimation.

A number of 45 patients in a cluster was estimated to provide a 90% power to identify the clinical outcome of interest (20% strain value difference between groups), with a significance level (alpha) of 0.05 (two-tailed)^10^.

This case-control matched study was made between October 2017 and February 2019.

### Body mass index (BMI)

Body mass index was calculated by the formula weight (kg)/height (m^2^). Obesity was contemplated when BMI exceeded >+2 SD for sex and age based reference values.

### Blood pressure (BP)

Blood pressure was measured with an appropriate cuff in supine position in the Endocrinology consult. Patients with hypertension (systolic and/or diastolic BP over the 90^th^ percentile for sex and height) were excluded off the study.

### Laboratory findings

In all obese children a complete analytic was obtained: fasting blood glucose, glycosilated haemoglobin, homeostatic model assessment of insulin resistance (HOMA-IR), lipidic profile and calcium phosphate metabolism.

### Cardiovascular evaluation

Including personal interview (functional class, physical activity level, sleep quality, familiar and personal cardiological backgrounds), and physical examination (cardiopulmonary auscultation, pulse palpation).

A functional echocardiography was performed to all patients both standard (left ventricular end-diastolic diameter-LVEDD, ejection fraction-EF) and functional parameters (tricuspid annulus plane systolic excursion-TAPSE, estimation of pulmonary pressure for assesing the right ventricle-RV function; longitudinal global and regional strain, pulsed-wave Doppler and tissue-Doppler for assesing the left one).

This study was made by only one observer using a Philips EpiQ team provided with Premium Ultrasound System technology.

TAPSE is a standard procedure to take the measurement of the distance of systolic excursion of the RV annular segment along its longitudinal plane, from a standard apical 4-chamber window, and represents longitudinal function of the RV^11^. Peak tricuspid regurgitation (TR) velocity was evaluated to calculate the RV-right atrial gradient and estimate pulmonary artery systolic pressure (PAPS)^12^. TAPSE/PASP ratio is a substantial prognostic parameter in patients with pulmonary hypertension and is related with hemodynamic and functional class^13^. This ratio was subsequently calculated.

Left ventricular determinations were executed in concordance to the recommendations of the American Society of Echocardiography^14^. Ejection fraction was assessed using the Teichholz routine.

Diastolic function was analysed by the mitral inflow, pulsed-wave Doppler (PWD) and tissue-Doppler imaging (TDI). TDI was obtained at the lateral margin of the mitral annulus and the velocity of the e’ wave was measured. Principal values of lateral e’ wave was captured and the E/e’ratio was descended from this mean value^15^.

The images obtained from standard apical 4-chamber, 3-chamber and 2-chamber views were then processed (echoPAC) to calculate the end-systolic longitudinal global strain (LGS) according with the consensus document of the European Association of Cardiovascular Imaging^16^.

### Statistical analysis

Descriptive analyzes of all the variables were carried out separately, using tables of absolute relative frequencies in the case of quantitative variables, and by mean ± SD, in the case of continuous quantitative variables. The Kolmogorov-Smirnov test was used to assess the normal distribution of the data.

The bivariate analysis was performed using the chi-square test for the comparison of proportions and those of Student Fisher-t to compare two means. Pearson’s linear correlation coefficient analysis was used to check the positive or negative correlation between BMI, laboratory and echocardiography parameters.

p< 0,05 was considered statistically significant. All the statistical tests were bilateral, considering 95% confidence level. The calculations was made with the help of the statistical package SPSS 22.0.

## Results

The study included 52 obese children (25 boys and 27 girls, mean age 10,98 ± 1,63 years) and 52 controls (32 boys and 20 girls, mean age 11,04 ± 2,91 years) referred to cardiologist by other reasons (murmur, thoracic pain or syncope) with structurally normal hearts.

Mean BMI in obese children was 28,37 ± 3,13 kg/m^2^, SD +2,54 ± 0,89.

Mean SBP and DBP in obese children was 117,64 ± 12,90 mmHg and 67,52 ± 9,85 mmHg, respectively. One patient with systolic blood pressure (SBP) 165 mmHg and diastolic blood pressure (DBP) 85 mmHg confirmed by Ambulatory Measurement of Blood Pressure was excluded from the study.

Fourteen obese children presented limit total cholesterol 171-199 mg/dL and only one hypercolesterolaemia of 223 mg/dL. Four patients had levels of HDL fewer than 35 mg/dL. Hypertriglyceridaemia > 150 mg/dL was seen in five obese subjets.

14 patients (27%) presented vitamin D deficience.

17 patients (32,7%) were still not in puberty (Tanner <2).

The mean insulin value was 25,68 ± 12,05 μu/mL and HOMA-IR was 5,34 ± 2,39 in the obese subjects. Insulin resistance (HOMA-IR > 2,5) was identified in 30 of them (57%).

The standard parameters between the both groups were compared in table 1. The thickness of interventricular septum-IVSd (p= 0,026), posterior wall-LPWd (p=0,018) and LVEDD (p= 0,005) were significantly greater in obese children, revealing cardiomegaly and hypertrophy in standard echocardiography.

**Table 1.**
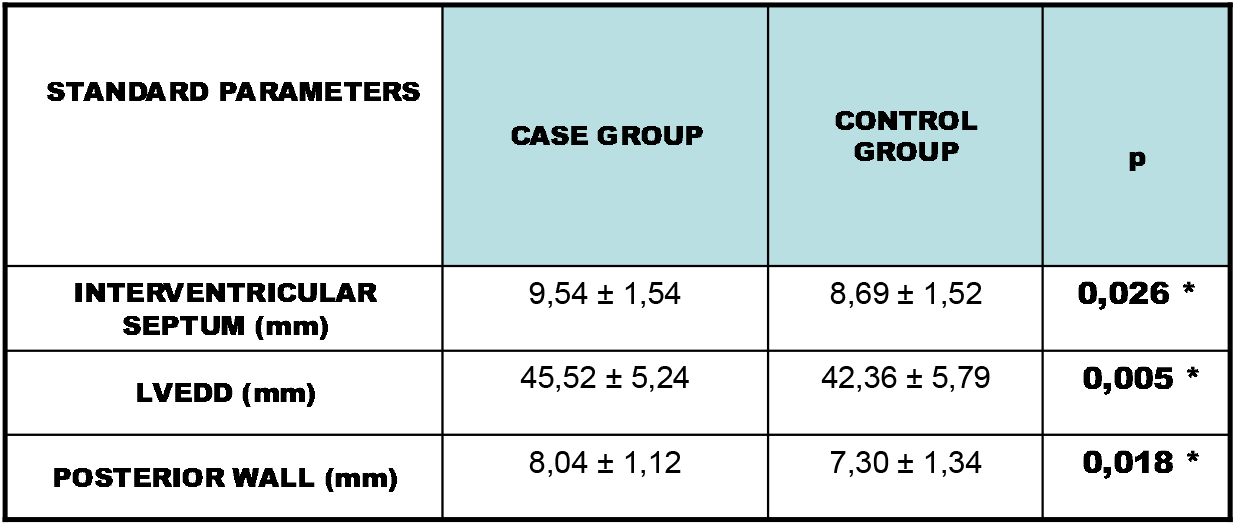

| STANDARD PARAMETERS | CASE GROUP | CONTROL GROUP | p |
| --- | --- | --- | --- |
| INTERVENTRICULAR SEPTUM (mm) | 9,54 ± 1,54 | 8,69 ± 1,52 | **0,026 \*** |
| LVEDD (mm) | 45,52 ± 5,24 | 42,36 ± 5,79 | **0,005 \*** |
| POSTERIOR WALL (mm) | 8,04 ± 1,12 | 7,30 ± 1,34 | **0,018 \*** |

Functional parameters are represented in **Table 2**. Pulmonary pressure was significantly higher in the case group (p= 0,002).

**Table 2.**

| FUNCTIONAL PARAMETERS | CASE GROUP | CONTROL GROUP | p |
| --- | --- | --- | --- |
| TAPSE (mm) | 23,02 ± 3,26 | 22,20 ± 3,68 | 0,242 |
| PULMONARY PRESSURE (mmHg) | 27,79 ± 6,10 | 24,31 ± 4,58 | **0,002 \*** |
| TAPSE/PAPS | 0,86 ± 0,19 | 0,93 ± 0,20 | 0,124 |
| LONGITUDINAL GLOBAL STRAIN (%) | -15,90 ± 3,84 | -19,44 ± 5,75 | **0,001 \*** |
| E/A | 1,61 ± 0,33 | 1,72 ± 0,32 | 0,157 |
| E/e' | 5,97 ± 1,21 | 6,28 ± 1,83 | 0,399 |

Longitudinal end-systolic global strain (LGS) was poor in obese children in comparison with controls, throwing a significant systolic subclinic dysfunction (p=0,001). Diastolic function assesed by PWD and TDI was not different between the two groups.

In **Figure 1** is represented the global and regional strain in a control patient (“bull’s eye like”). LGS is derived from speckle tracking and analyzed by post-processing of apical images of the left ventricle (LV). Longitudinal shortening is expressed like a percentage.

**Figure 1.**
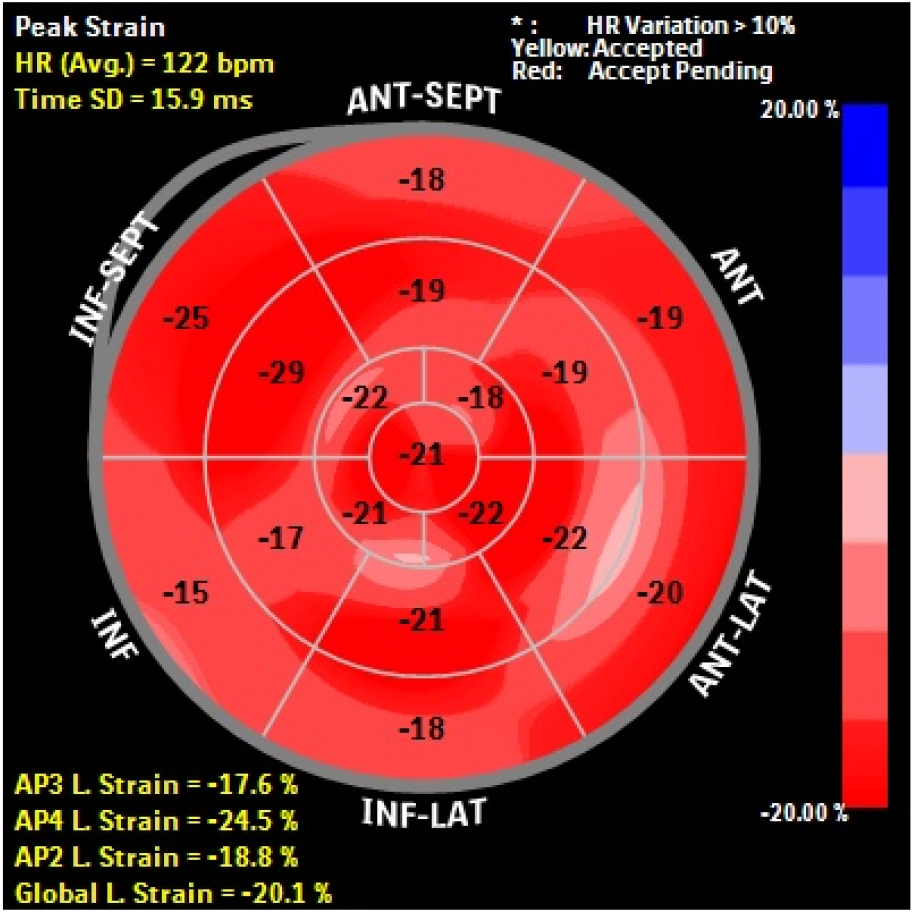

The apical image of LV in a obese patient is showed in **Figure 2**. Note that inferior, anterior and anteroseptal wall present the worst changes in length in proportion to baseline. This is a relatively common find in our study but normal values of regional strain are not still validated in pediatric population.

**Figure 2.**
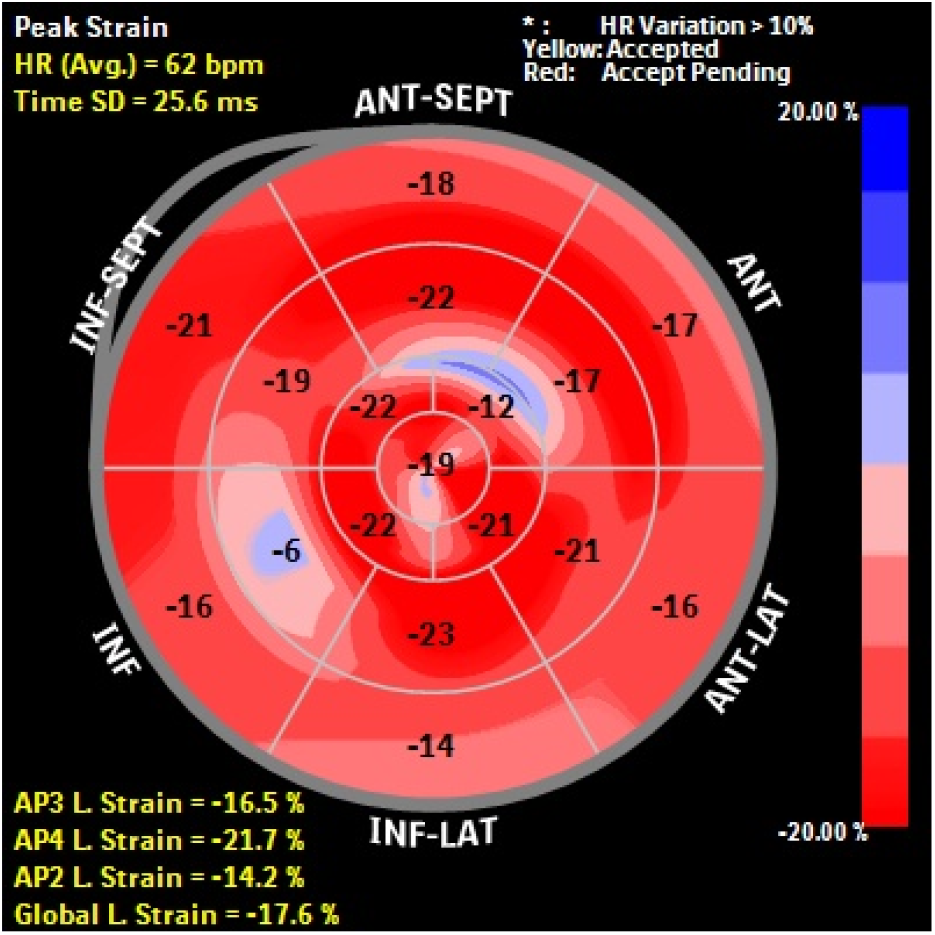

Correlation between IMC and some laboratory data with ecocardiographic parameters are represented in **Table 3**. Those marked in bold are significant at p<0,05.

**Table 3.**

|  | **BMI**<br>**(kg/m<sup>2</sup>)** | **HOMA-IR** | **TOTAL**<br>**CHOLESTEROL**<br>**(mg/dL)** | **TRIGLYCERIDES**<br>**(mg/dL)** |
| --- | --- | --- | --- | --- |
| **INTERVENTRICULAR**<br>**SEPTUM (mm)** | **0,346** | 0,347 | -0,039 | 0,309 |
| **LVEDD (mm)** | **0,478** | 0,328 | -0,170 | -0,029 |
| **POSTERIOR WALL (mm)** | **0,358** | **0,486** | -0,158 | 0,008 |
| **TAPSE (mm)** | -0,022 | -0,264 | -0,208 | -0,249 |
| **PULMONARY PRESSURE**<br>**(mmHg)** | **0,321** | 0,112 | **0,330** | **0,460** |
| **TAPSE/PASP** | -0,217 | 0,355 | **-0,419** | **-0,503** |
| **LONGITUDINAL GLOBAL**<br>**STRAIN (%)** | **0,533** | 0,224 | -0,064 | 0,005 |
| **E/A** | -0,136 | **-0,458** | **-0,297** | **-0,402** |
| **E/e'** | -0,035 | 0,130 | 0,055 | -0,124 |

There was positive correlation between BMI and all standard parameters (IVSd, r 0,346, p=0,014; LVEDD, r 0,478, p=0,000; PWd, r 0,358, p=0,013), and between HOMA-IR with PWd (r 0,486, p=0,012).

Pulmonary pressure correlated positively with BMI (r 0,321, p=0,017), total cholesterol (r 0,330 p=0,033) and triglycerides levels (r 0,460, p=0,005). However TAPSE/PAPS ratio correlated negatively with total cholesterol (r −0,419, p=0,006) and triglycerides levels (r −0,503, p=0,002).

We found significative positive relation between BMI and LGS (r 0,533, p=0,000). HOMA-IR (r −0,458, p=0,006), cholesterol (r −0,297, p=0,048) and triglycerides levels (r −0,402, p=0,011) correlated negatively with diastolic function of left ventricle estimated by E/A ratio with PWD. This correlation was not extensive to E/e’assesed by DTI.

Multiple linear regression analysis using the backwards method was performed. The dependent variable PWd was significantly correlated with HOMA-IR (β 0,486, p=0,012). Pulmonary pressure correlated positively with BMI (β 0,458, p=0,003) and total cholesterol (β 0,347, p=0,023). TAPSE/PASP ratio correlated negatively with triglycerides levels (β −0,402, p=0,014). Diastolic function was affected by HOMA-IR (β −0,375, p=0,016) and triglycerides levels (β −0,375 p=0,024).

## Discusion

Obesity in children has become an authentic pandemic in developed countries. There are different criteria for defining overweight and obesity, to facilite the comparison between different studies and countries, taking into account the ethnic diversity and avoiding underestimation that involves comparing with populations in which obesity already existed.The criteria most commonly approved by the international scientific society are those recomended by International Obesity Task Force (IOTF)^17^ (cutoff points for childhood overweight or obesity particular to each BMI value and sex, using the reference population specified by the IOTF) and those proposed by the World Health Organization (WHO)^18^ (overweight, specific body mass index (BMI) values agreement to age and sex, >+1 standard deviation (SD) in the reference population characterised by the WHO; obesity: sex-and age-specific BMI values, >+2 SD). In our study we have chosen the WHO criteria.

Enkid study (1998-2000)^19^ has been the reference in obesity in children in Spain for many years, but it’s outdated.

The two most recent national studies used direct height and weight measurement: ALADINO^20^ (Diet, physical Activity, Childhood Development and Obesity) study with children aged 6 to 9.9 years old; and a cross-sectional, observational study composed of children and adolescents of both sexes between 8 to 17 years old residing in family homes in mainland Spain^2^. The prevalence of overweight and obesity in this study is 26.0% and 12.6%, respectively. In our region the prevalence of obesity is quite higher (18,5%)^21^.

Among obese children, based on our results, standard echocardiography did not show anomalies in the global and segmental contractility. However, LVEDD, interventricular septum (IVSd) and posterior wall of left ventricle (LPWd) measured in diastole were significantly higher in obese group in comparison with controls (p =0,005, p=0,026 and p=0,018, respectively). A subgroup of obese children could present LV remodelling characterized by increased volume and wall thickness. These findings are superposable to Binnetoglu et al’s^22^.

In healthy children, the mean LGS value is −20,5%^23^. In our study, obese children compared with controls had lower 2D STE-derived LV LGS (−15,90 ± 3,84% vs 19,44 ± 5,75%, p=0,001). LGS in our study was not affected by sex or age.

Similar observations of decreased longitudinal and circumferential strain in children with obesity were concluded by Kulkarni et al^24^. These findings are extensible to radial strain and early diastolic strain rate values^25^. Segmental analysis of the LV could provide in the future subtle markers for the emergence of future obesity-related cardiac disease^26^.

Current guidelines recommend pulmonary hypertension screening in patients with severe obstructive sleep apnea. Prevalence of pulmonary hypertension in pediatric patients with obstructive sleep apnea is low and none of the patients with pulmonary hypertension had severe obstructive sleep apnea in a recent study over 163 patients ^27^. According to this in our study we found significative higher pulmonary pressure in obese children, but no one had pulmonary hypertension (PAPs over 40 mmHg).

Normal TAPSE values including z-scores based on patient age have been established in pediatric patients^11^. The TAPSE/PASP ratio has recently been reported as an independent prognostic parameter in heart failure. When stratified by tertiles, patients in the low tertile showed significantly compromised hemodynamic, functional and echocardiographic status^13^. This index may be a step forward a more efficient RV function evaluation. In our sample TAPSE/PAPS was significantly negative correlated with triglycerides levels (β −0,402, p=0,014).

Diastolic function of obese participants was assessed by pulsed-wave Doppler (PWD) and TDI. We didn’t find a statistically significant difference that could indicate impaired LV filling like Gandhi et al^28^. However we found relation between HOMA-IR and triglycerides levels, correlated negatively with diastolic function of left ventricle estimated by E/A ratio with PWD. In previous studies LV early diastolic strain rate lower in hypercholesterolemic children compared with normals is also described^29^.

And now the question is, can we take an active attitude with these patients that have already remodeling and some index of RV and LV function altered? Gores et al^30^ describe an improvement in RV function assessed by TAPSE in a group of overweight patients participating in vigorous 3-month football training. We think next interventions could follow this line and select between the obese patients those that could beneficiate from personalized training and enter in a program of cardiac rehabilitation.

### Limitations

The statistical power of the analysis is limited by the small sample size.

There wasn’t positive correlation between HOMA-IR, cholesterol, triglycerides and remodelling of left ventricle^31,32^. One reason could be that we didn’t estimate the LV mass index.

We didn’t find relation between HOMA-IR, total cholesterol and triglycerides levels with LGS. Perhaps the view selection, defining end-systole or tracing the myocardium was not the most adequate in some patients that had bad window.

## Conclusion

Remodelling of LV and subclinic systolic dysfunction could be relatively common in asymptomatic patients with obesity and easily detected by functional echocardiography including TAPSE, PAPS and 2D-STE.

We think that is necessary to perform an standardized cardiovascular evaluation in obese children for early identification of subclinical dysfunction especially in those with insulin resistance and dyslipidemia

## Data Availability

The data that support the findings of this study are available from the corresponding autor, upon reasonable request

